# Public relations strategies employed by healthcare organizations to address vaccine hesitancy: the case of the Ghana Health Service

**DOI:** 10.1101/2022.08.10.22278623

**Authors:** Ruth McCarthy, Emmanuel Timmy Donkoh, Dominic De Graft Arthur, Edward Tieru Dassah, Kwame Ofori Boadu, John Ekow Otoo, Ivy Wina Ofori Boadu, Samuel Fosu-Gyasi

**Author notes:** Correspondence should be addressed to Emmanuel Timmy Donkoh.

## Abstract

**Background:** Strategies for developing and advancing good public relations can be recognized in nearly all fields of life without making an exception for the healthcare industry. In the wake of the COVID-19 pandemic, matters of public health have gathered more force. The importance of effective public relations for improving healthcare is highlighted by the position that immediate access to reliable health information should be the hallmark of a just society. However, the strategies available for addressing major threats to the uptake of public health services such as mass vaccination campaigns are not properly studied and documented in the Ghanaian context.

**Methods:** Semi-structured interviews were conducted with officials of the Ghana Health Service (GHS). Participants were recruited through purposive sampling. Data collected included demographic characteristics, perspectives on public relations strategies used in the past year to improve vaccine uptake as well as successes and pitfalls. Thematic content analysis was performed on data collected using the NVIVO software.

**Results:** Healthcare workers perceived vaccine hesitancy to be a threat with the potential to undermine an important strategic organizational goal related to COVID-19 illness. A combination of informative, motivational, persuasive and coercive public relations strategies was employed by the Ghana Health Service to address the challenge of vaccine hesitancy. These strategies were deployed across both traditional (print, radio, TV) and emerging/social media networks. Officials were optimistic that the strategies would produce results but were uncertain whether they could attribute current successes or failures to the PR strategies used.

**Conclusion:** Since the onset of the COVID-19 pandemic, several public relations strategies have been evidently employed by the Ghana Health Service to address vaccine hesitancy. The nature of the audience and PR strategies employed suggests that the effect of these strategies may be short-lived unless they are constantly reinforced by the GHS.

## Introduction

The development of vaccines and universal vaccination programs have been significant in lowering the burden of infectious diseases and the number of related deaths (1). In the past, infectious diseases that are now easily preventable were responsible for several deaths. The success of vaccines have led many to downplay the danger posed by emerging infectious diseases and the potential loss of lives in the absence of effective vaccines (2). There is even growing doubt and negative speculation about the beneficial outcomes and efficacy of vaccination in improving the health of populations (3). These suspicions have led to vaccine hesitancy (4) and dwindling vaccination coverage (5).

For an immunization program to work effectively as a public health strategy, healthcare staff must be able to implement it at scale (6, 7). The mere availability of safe and efficacious vaccines is only one side of the coin; high rates of acceptance and utilization by members of the larger population is critical to driving intended benefit (7). Nearly universal vaccination coverage is essential (8), to guarantee protection for individuals, and also for attaining the drastic reduction in transmission that is typically termed “herd immunity” (9).

Unlike many other pharmacological interventions, public confidence in vaccination is at an all-time low following the COVID-19 pandemic; influenced by unhealthy suspicion of the safety and effectiveness of vaccines, skepticism about the interests of health workers and healthcare organizations and their products, and of the policies of the scientific establishment. At the peak of the COVID-19 pandemic, efforts by governments around the world to rapidly fund, develop and distribute vaccines with emergency use authorization from the World Health Organization (WHO) led to fears about vaccine quality and safety (10). Aversion to vaccines may by mediated by health beliefs. Ironically, the far-reaching success of vaccination may in itself be a catalyst for anti-vaccine sentiment by lowering perception of disease risk and severity. Factors sustaining aversion to COVID-19 vaccines resemble well-documented general anti-vaccine sentiment. Broadly, these can be classified into vaccine-related attributes, vaccine-related attitudes and beliefs, and the political environment (11).

The devastating impact of infectious diseases such as Ebola and COVID-19 on healthcare systems with attendant lock-downs and travel restrictions in several African countries highlight the importance of vaccines. However, there is no guarantee that once vaccines are available, they will be acceptable to the population. Available data on public attitudes toward vaccines indicate substantial vaccine complacency (12). To overcome this scenario, a multi-pronged evidence-based approach is required to instigate behavior change and address vaccine hesitancy. Undoubtedly, the best examples of public relations (PR) need to be considered by healthcare organizations to leverage upon credible and transparent political leadership in order to drive a robust health delivery to redress the “infodemic” fueling vaccine hesitancy (13). We sought to explore effective PR strategies used by health care professionals to address vaccine hesitancy in the country.

## Materials and Methods

### Study design and setting

A qualitative explorative descriptive design was employed as very little is presently known about the PR strategies employed by healthcare managers to promote vaccine compliance in the Ghanaian context. The study was conducted among officials of the Ghana Health Service (GHS) from the Bono Region of Ghana. The GHS is an administratively autonomous organization established by law (Act 525 of 1996). It is vested with responsibility to implement national policies formulated by the Ministry of Health (MOH) through the Ghana Health Service Council which serves as the governing board. Following the reorganization of the MOH and healthcare administration in the country, it was recognized at the time that the growing level of managerial responsibility that was delegated to districts hospitals required a new organizational paradigm to oversee. The formation of the GHS comes as one of the vital strategies recognized by the Health Sector Reform of the nineties, which were guided by the Medium-Term Health Strategy in the pursuit of establishing a more equitable, efficient, accessible and responsive health care system.

The GHS is the sole implementing authority of health policy in Ghana by providing and managing comprehensive and accessible health service with special attention for primary health care at regional, district and sub-district levels. Promoting health, mode of healthy living and good health habits by people is seen as a key function. The GHS has 16 Regional Health Directorates, with one in reach of the 16 regions of the country. Each region also has district health directorates. The PR responsibilities of the GHS are a function of the office of the Director General. However, as a result of the stratified nature of health administration, district and regional directors of health also play a significant role in engaging the public.

### Participants and recruitment

Key informants or officials with the appropriate managerial experience were purposively selected to provide feedback relevant to the phenomenon under study (14, 15). At the regional and district offices of the GHS in the Bono Region, the general purpose of the study was discussed with a large group of selected officials and those who consented were scheduled for interviews. Selected officials or key informants were provided information about the rationale for the study and issues regarding ethical conduct of research such as voluntary participation, potential risks and respect for privacy when reporting all findings were agreed in advance. Interviews were conducted through an electronic instrument or in-person as scheduled until data saturation was attained. Data saturation occurred when no new themes and subthemes emerged after conducting three consecutive interviews.

### Data Collection

Data for the study was collected from November to December, 2021. Participants were contacted in advance via telephone calls and text messages to remind them and confirm all interview arrangements. Participants were also required to complete a short online survey. Informed consent (including permission to digitally record the interview session) was documented before in-depth one-to-one interviews with managers and PR officers were conducted by a researcher with experience in qualitative research.

All interviews were opened with an exchange of pleasantries and general conversation to create rapport and to set respondents at ease to give feedback without inhibition or apprehension. Assurances were also given regarding the confidentiality of conversations and participant’s right to anonymity. This created a congenial environment for data collection with the aid of a semi-structured interview guide (S1 Appendix A.docx: Interview guide for in-depth interviews). Participant’s demographic data including gender, official designation and years of work-related experience were captured.

### Research Tool/Instrument

The study relied on a semi-structured interview guide to elicit information about (1) participants’ knowledge of the threat of vaccine hesitancy to the achievement of organizational goals (2) PR strategies employed by the organization to improve attitudes toward vaccinations (3) the impact of available PR options (4) contextual factors that promote the success or failure of these PR strategies. A consultative approach was followed to develop the instrument based on empirical literature and expert guidance. The instrument was pretested in a healthcare organization to elicit feedback on test reliability and validity for improvement. The interview guide can be found as a supplementary file (S1 Appendix A.docx: Interview guide for in-depth interviews).

### Data management and analyses

Data collected from in-depth interviews was subjected to the thematic content analysis procedure by Braun and Clarke(16) using NVIVO software version 10.0 (QSR International, MA, USA). The approach was useful for conducting a thorough analysis of the data in search of meaning (17). All coding of the data was done in duplicate in NVIVO and nodes or aggregates plotted into potential themes. Potential themes were discussed in a series of conferences as recommended until they were finalized by group consensus (16).

### Rigour

A qualitative study report is considered trustworthy when it is credible, transferable, dependable, and confirmable (18, 19). Credibility is used to examine the level to which the research results reflects the reality and is representative of the participants’ views (18, 20). The principal investigator checked with participants to fully understand ideas represented and adequately captured participants’ story lines to ensure credibility. Dependability refers to the quality of the integrated processes of data collection, data analysis, and theory generation that can be audited (20). This was achieved by a detailed description of the research methodology (recruitment process, data collection, data analysis) in line with the consolidated criteria for reporting qualitative research (COREQ) reporting guidelines (19, 21). Transferability refers to how the study results could be used in different areas and contexts (19). To ensure this, a comprehensive description of the study setting was provided. Confirmability was ensured by keeping records of the field notes and voice records.

### Ethical Issues

The study was approved by Committee for Human Ethics, the Institutional Review Board of the University of Energy and Natural Resources. The study was conducted in conformity with the requirements of ethical propriety: all participants were adequately informed about the study before indicating their desire to participate in recorded interactions and also provided written voluntary consent for data provided to be anonymized prior to dissemination.

## Results

### Demographic Characteristics

Altogether, 15 participants comprising 8 males and 7 females granted interviews for the study. These individuals were aged between 36 years and 69 years. The number of years of participants’ experience in a managerial capacity ranged from 2 to 10 years and all participants had obtained a minimum of undergraduate education.

### Main Findings

Four main themes emerged from the data analysis and these are represented in Table 1. These are a) Organizational goals and potential threats b) Knowledge of public relations strategies in use by organization c) Tools/Media employed for the realization of public relations strategies d) Anticipated impact of PR strategies. In addition to the main themes, a number of sub-themes were synthesized.

***Table 1: Summary of themes and sub-themes from the transcribed data***

### Perceived threat of vaccine misinformation to achievement of organizational goals

Vaccination of the public was considered to be an important public health target. The organization was in pursuit of a threshold phenomenon described by participants in technical language as “herd immunity”. Most participants considered this goal of their organization to be somewhat under threat insofar as large aspects of the public remained skeptical about the judicious and safe use of the vaccines.

> *Yes. Covid 19 is easily spread from one person to another. With the vaccine, transmission rate can be reduced so that the vulnerable will be protected (that’s the whole point of herd immunity) [Official 005; 4 years’ experience]*
>
> *Yes, Vaccine hesitancy is very much present among the general populace and this can adversely affect targets, hinder the achievement of herd immunity and increase the chances of being infected and its complications [Official 004, 5 years’ experience]*

In terms of how hesitancy constituted a threat, respondents anticipated that negative sentiments could delay the attainment of herd immunity.

> *Yes. It will lead to delays in achieving herd immunity [Official 010; 3 years’ experience]*
>
> *…any obstruction to increasing vaccine acceptance and coverage slows us down in attaining these targets [Official 013; 10 years’ experience]*

However, there were various obstacles to be overcome in order to successfully obtain this organizational goal. One such threat was non-adherence to vaccine advice:

> *Yes, vaccine hesitancy is a threat to the vaccination program. People refusing to vaccinate will prevent the achievement of the set coverage that will enable herd immunity and therefore the full benefit of vaccination to the communities and the country in general [Official 009; 2 years’ experience]*

A number of reasons could be attributed to the threat of hesitancy such as religious persuasion, misinformation, lack of proper orientation regarding the scientific basis of vaccines and the result of widespread misinformation across multiple social media channels.

> *This is as a result of myths and misconceptions about the side effects of the vaccines and other religious beliefs [Official 011; 8 years’ experience]*

In spite of the perceived threat of vaccine hesitancy to the organization’s goals, this was more of a perception than the prevalent reality. This compelled a few respondents to believe that vaccine hesitancy was not a threat at all to the organization’s ambitions.

> *No. Vaccine hesitancy is not primarily an issue. Most patients request for [the vaccine] voluntarily. The problem is the availability [Official 014; 6 years’ experience]*

However, participants agreed that these threats were of such a nature that they could be addressed with effective PR strategy. Respondents reported coming across individuals with beliefs ranging from the very entrenched to mere ‘hearsay’ about the harmful nature of vaccines. It was considered that to some extent, there was a real possibility that negative beliefs could be dispelled by offering accurate scientific information through credible officials and agents.

> *Vaccine hesitancy though can affect the expected target: [however,] the challenge can be addressed by public health personnel using the right approach and the best information [Official 008; 4 years’ experience]*
>
> *This is because people have a whole lot of negative ideas and philosophy about this vaccine. [Effective] PR work will help explain the issues better and also improve public sentiment so that more people will be willing to take the vaccine [Official 003; 8 years’ experience]*
>
> *Vaccine hesitancy is strongly influenced by misinformation*…*conspiracy theories over social media about the COVID vaccines are the main cause [Official 012; 5 years’ experience]*

The expert interviews also revealed the need for health organizations to promote PR practice and to engage the public by confronting the new wave of misinformation and adapting to the speed of information spread through new and emerging media.

> *The EPI [Expanded Programme on Immunization] was successful because of PR. However, that was easier in those times because most information came from National TV and radio stations. So, it is an issue of adapting PR strategies to modern times [Official 004; 5 years’ experience]*
>
> *A target-specific public relations strategy can help dispel some of the myths peddled by anti-vaccine campaigners and promote a more detailed health education on the advantages of vaccines [Official 006; 5 years’ experience]*

### Public relations strategies used by the GHS to promote vaccine acceptance

Since the start of the pandemic, public health managers at the GHS have devised various strategies to engage their publics and generate sympathy towards their organization’s objectives. The present study identified a number of themes reflecting elements of PR strategy embedded within the messages encoded by officials of the service and broadcast to the public. The elements of PR strategy discovered from interactions with key personnel of the GHS are outlined.

### Informative PR strategy

The Service proactively employed a communication strategy to make as much public information available as possible through their officers (Figure 1). Messages were broadcast on radio and television in the local languages and in English. Also, regular press briefings were held led by the Presidential Task Force on COVID-19 which was supported by the GHS. After each press briefing, the Minister of Health followed up with a meet-the-press session to elaborate on pertinent issues that needed clarification. The “open door” strategy where journalists could ask questions and get feedback from the Minister of Health and the leadership of the Health Service encourages radio and television stations nationwide to dedicate airtime to broadcast the briefing sessions and helped the GHS to set the tone for public discussion with authoritative information throughout the COVID 19 pandemic.

**Figure 1:**
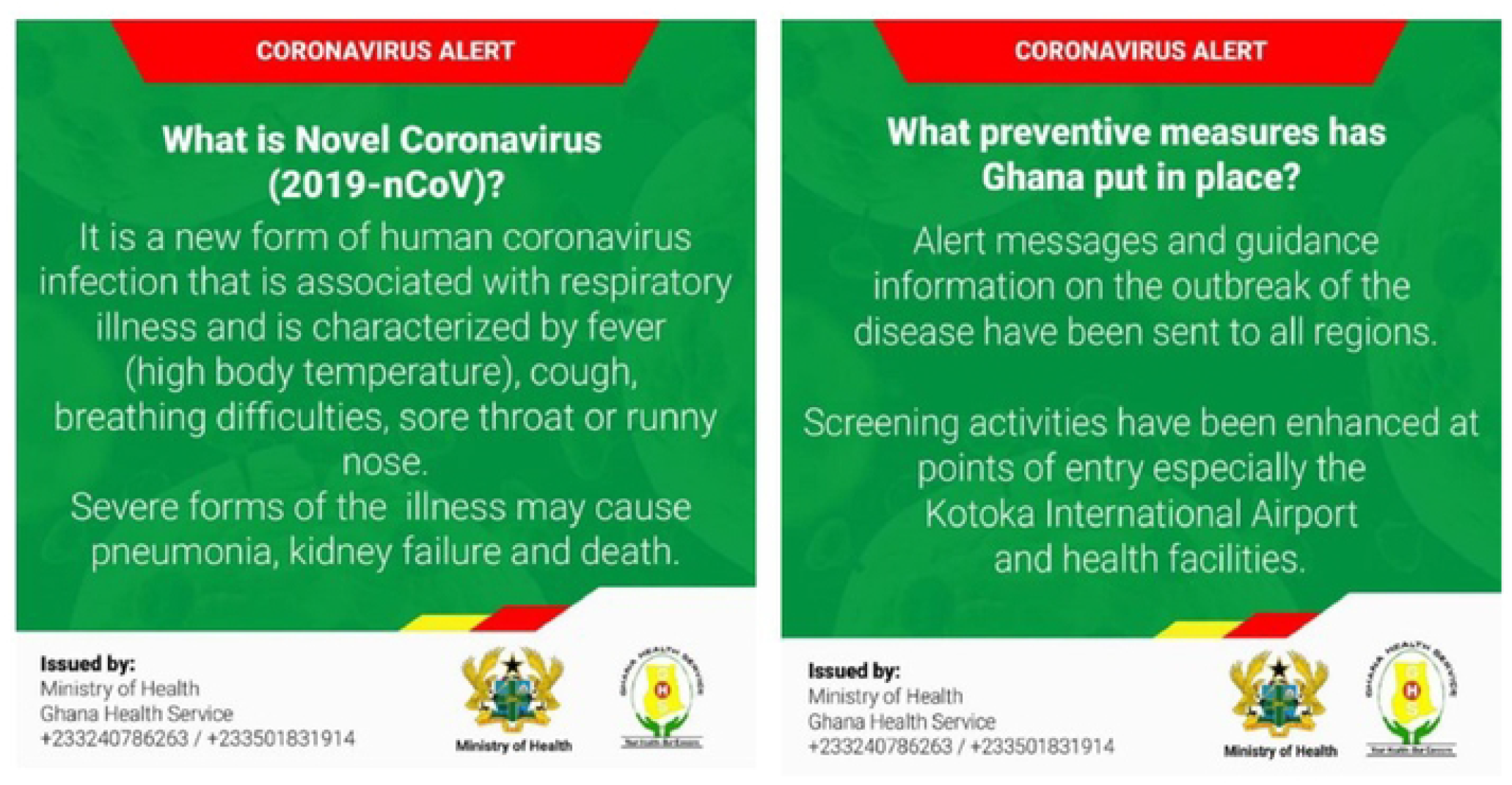
Infographics depicting informative public relations strategy.

> *We sent our men to the radio stations to educate the public on the dangers of refusing to accept the vaccines. We told them to stress the need for radio stations to talk to the experts. You know charlatans try to take advantage of people by posing as knowledgeable individuals. I think to some extent the strategy worked [Official 001; 3 years’ experience]*

From the early days of the COVID-19 pandemic, the GHS has relied on electronic and print media to provide education to people on the importance of taking the vaccine. Through the Disease Control Unit, the GHS has produced and circulated informative educational material. Samples of these are shown in Figure 1. Some officials noted:

> *At almost all of our workshops, the need for more education was constantly stressed. We on the inside had access to vital information from the WHO: we attended webinars and several zoom meetings. But what about the populace? Who was organizing webinars for them? So, we felt that we needed to put out as much education as possible [Official 011; 8 years’ experience]*
>
> *We always felt that education was our best strategy to fight this [epidemic of misinformation]. Basically, some informative flyers were produced and circulated to our facilities, market places, and on social media [Official 007; 6 years’ experience]*

Additionally, a COVID-19 dashboard was launched to provide near real-time information about the rising number of cases of COVID-19. Throughout the pandemic, this dashboard was considered the most authoritative source of information and was mostly relied upon during official briefings.

> *I heard many conspiracy theories about Governments falsifying case numbers to impose lockdowns. But I think that here [in Ghana], the figures we put out on the dashboard has helped a lot… [Official 014; 6 years’ experience]*

### Facilitative PR strategy

The GHS deployed elegant vaccine-related communication to create awareness about vaccination schedules, sites and protocols. These infographics were meant to be circulated widely on social media pages to provide reliable information about vaccination schedules and thereby facilitate uptake of vaccines by citizens (Figure 2).

**Figure 2:**
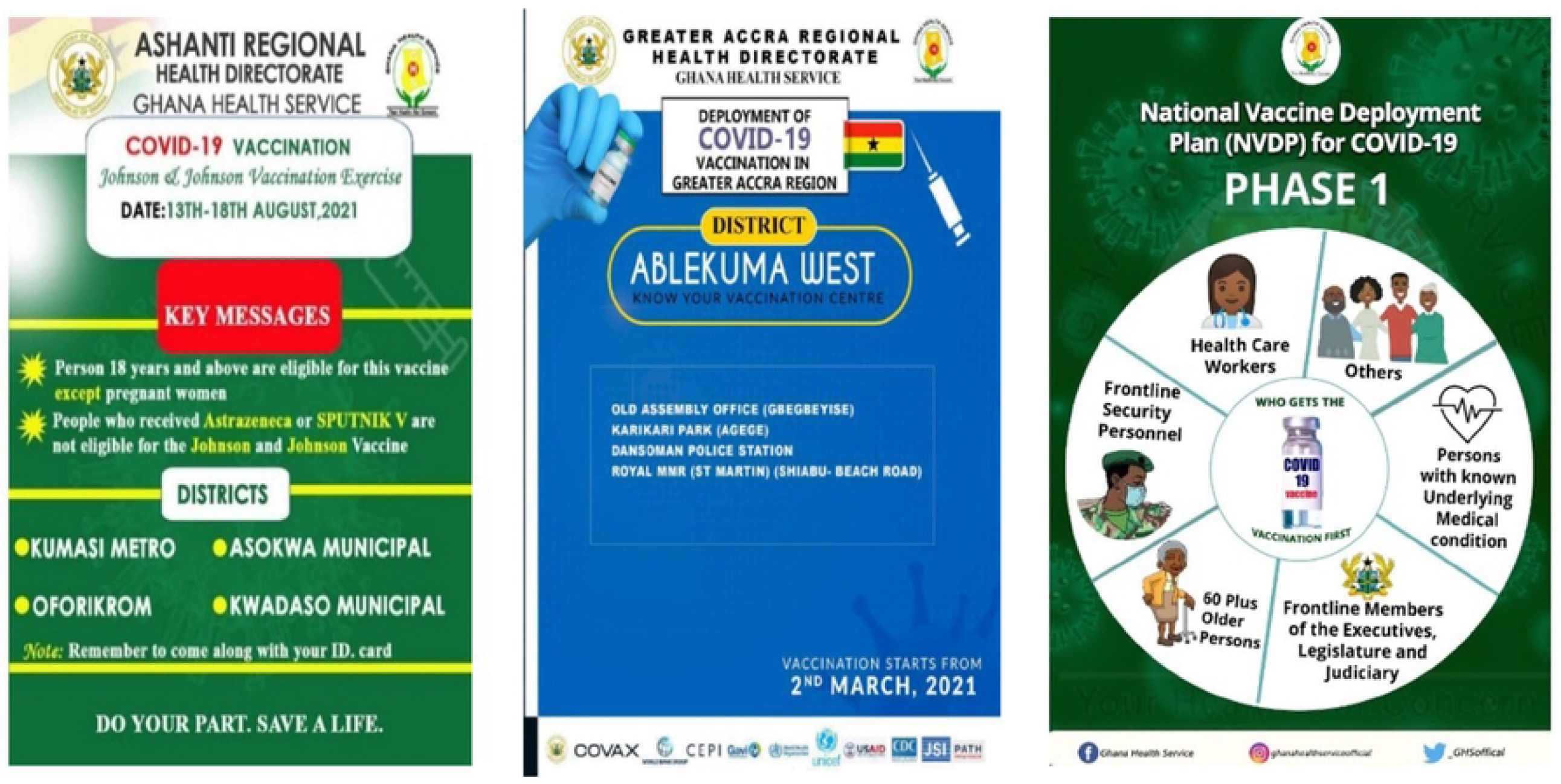
Infographics depicting facilitative public relations strategy.

> *If you want the people to buy into the exercise, you don’t want them to be frustrated by a lack of information that allows them to act independently… these are traders, office workers and generally busy people… We situated vaccination sites close to them and we tried to communicate that ahead of time [Official 015; 2 years’ experience]*

### Persuasive PR strategy

Another public strategy employed by the GHS was the use of press conferences with messaging calculatedly crafted to appeal to the public’s values and emotions and to instigate public sentiment for vaccination.

> *One of the things that we also did was to engage high-profile individuals and empower them to spearhead our communication. In this strategy, the President of Ghana readily comes to mind. In fact, this strategy was so useful it attracted international commendation [Official 005; 4 years’ experience]*
>
> *The use of opinion leaders, organized groups, and influential persons to champion the course of the COVID-19 vaccination was a vital strategy for us in addition to the use of social mobilization and risk communication messages to increase the uptake of the COVID-19 vaccination [Official 009; 2 years’ experience]*

The service also arranged for the first dose of vaccines in the country to be given to prominent chiefs, political figures and prominent civil society actors to persuade the citizens of their safety and avoid hesitation.

### Coercive PR strategy

A coercive strategy has also been employed by the Service. Although no vaccine mandates have been issued in Ghana at the time of this report, this does not mean that it has escaped officials from the Health Service. Repeatedly, high-ranking officials have been on record attempting to convince members of the public through messages that hint at the imminent imposition of restrictions at border posts and social gatherings.

> *Before the Christmas festivities, we secured enough vaccines for the population. We wanted as many people as possible to get the jab. Therefore, I am not surprised that there were hints that without getting vaccinated, for instance, you can’t attend public events [Official 013; 10 years’ experience]*

More commonly, the coercive strategy has been used by officials of the GHS when referring to travel requirements for passengers to produce evidence of a negative polymerase chain reaction (PCR) test result within 72 hours of boarding at international airports.

### Motivational PR strategy

In order to maintain a positive internal sentiment about vaccination within the organization, the GHS employed various communication strategies to keep health workers motivated. Health workers were depicted in several ways as “front-line”, “at-risk”, “heroes” and the like to keep them as motivated as possible. These adjectives were meant to court public sympathy, promote organizational values and ultimately prevent and reduce hesitation about vaccination. In addition to motivating language descriptors, health workers received regular testing services at no cost, donations of personal protective equipment, access to paid off days, priority access to vaccines and monetary reward from the state for their work in helping to promote health. These actions were constantly mentioned during regular press briefings organized by the Service and the Presidency.

> *You know [pause] healthcare workers are role-models for friends, family and society at large. We all saw viral videos of healthcare workers in other countries, and even in Ghana, calling on the public to abstain from COVID-19 vaccinations. So, from the very start steps were taken to motivate our staff to step forward to be vaccinated and encourage others to do same [Official 004; 5 years’ experience]*

### Tools/Media employed for the realization of public relations strategies

Tools employed by the Service for the realization of organizational goals related to vaccines ranged from stakeholder engagements from the planning to the implementation stages, public education on COVID-19 vaccination through community durbars and use of community information centres, setting up mobile vaccination posts at health facilities and vantage points, advocacy meetings with opinion leaders and social groups and public health emergency committee meetings to provide feedback on all COVID-19 interventions.

> *We can also talk about infographics on TV, on social media, and in hospitals, all these tools were employed to drive strategic messages [Official 001; 3 years’ experience]*

#### Public engagement through traditional media

Traditional media channels were seen as important outlets by officials of the GHS for the dissemination of information and public engagement. Both national and private radio and television channels were used to broadcast press briefings, interviews, and soundbites and generally mobilize community members.

> *Some of the strategies employed by the GHS include 1. Continuous social mobilization using different media outlets to educate people on the vaccination process and the benefits thereof. 2. Continuous press releases on the epidemiology of the COVID-19 outbreak is one strategy that is to help with vaccine hesitancy 3. Use of role models in the society such as the president, vice president and their spouses to take the jab on national television, among others. [Official 003; 8 years’ experience]*

Participants felt that although the presence of the GHS on social media was a show of the Service’s willingness to evolve to keep up with the changing demands of the times, the traditional media such as radio and television was still essential for generating the right public sentiment because of the tendency for social media to be used to carry misleading information orchestrated by pranksters and other groups opposed to vaccination and other medical products.

#### Public engagement through new and emerging media

The GHS also embraces new and emerging channels such as websites, Instagram, Twitter and Facebook to promote communication targeted at improving vaccine behaviour. These channels convey communication crafted with the aforementioned PR strategies to a wide range of audiences who may otherwise be missed by traditional media such as adolescents and young adults.

> *First of all, the Ghana Health Service has a dedicated website where all official information concerning COVID-19 and vaccines is found. A COVID-19 helpline was announced last year as well: this is a 24-hour open line that people can access to have a professional discuss their misgivings about the vaccines. [Official 005; 4 years’ experience]*

#### Clinic/Facility engagements

As a complementary measure, the GHS also engaged members of the public who visited healthcare facilities with regular educative campaigns at out-patient departments (OPDs). These interactions between health workers and facility patrons could offer several advantages.

> *Forums are organized where the general public is allowed to ask questions and share their experiences related to the COVID vaccine and the vaccination process. [Official 015; 2 years’ experience]*
>
> *Education and testimonials are a major feature of our PR strategy. So, we do regular education at our OPD to sensitize people about vaccinations and to diffuse common myths. Also, testimonials of people who have taken it are brought, especially those who are popular and have integrity, so that others will know there is nothing to fear. [Official 012; 5 years’ experience]*

### Expected impact of PR strategies

The impact of the public relations strategy employed by the GHS can be described as positive overall. Participants felt strongly that the strategies implemented by the Service could play an important factor in preempting crisis levels of hesitancy by making more information available and evoking a cooperative public disposition.

#### Winning the information war with readily accessible credible information

> *People now have information that wasn’t available initially. All the unanswered questions about vaccines are being handled. [Official 001; 3 years’ experience]*
>
> *When there’s an official website or credible source, we can resort to resolving conspiracies and arguments with information from these places. [Official 006; 5 years’ experience]*

#### Increased uptake of services

Officials also spoke out about the impact of public relations practice and the success of interventions in Ghana

> *It has been positive because the application of the above PR [strategies] has led to an increase in the uptake of the vaccines and people are more committed to taking the vaccines [Official 008; 4 years’ experience]*
>
> *The above-listed PR strategies have helped [to] educate the people, cleared misconceptions and sustained confidence in COVID vaccination. [Official 003;8 years’ experience]*

#### Some amount of skepticism still prevails

Some participants were careful not to ascribe much success to the PR strategies employed citing entrenched beliefs and potential for complacency on the part of agents of the organization.

> *Not much. From my personal view, it’s only when close contacts to an individual who is hesitant contract the disease that perceptions change, no matter the PR strategy used. [Official 014; 6 years’ experience]*
>
> *The response has been good so far. Just that there is more work to be done. [Official 015; 2 years’ experience]*.

### Contextual factors that can account for the success or failure of these PR strategies

Participants gave their impressions about some contextual factors that in their opinion could promote or undermine the RR strategies discovered in the work. Key contextual factors that were identified to be relevant for the success of organizational PR regarding the threat of vaccine hesitancy were the need to build relationships of trust with community members over time, recruiting influential community figures to not only carry information to the community but also to mediate the relationship between the community and the organization. These influential persons are typically religious leaders, opinion leaders, market queens and elected representatives at the district assembly.

> *Strong social networks in the communities and trust in influential persons [can make all the difference]. [Official 009; 2 years’ experience]*
>
> *…the success of any PR strategy depends on how well-informed the PR person is and how well the information available to him is being delivered to the audience. However, if the information is not well delivered, the strategy will fail [Official 010; 3 years’ experience]*.
>
> *A key thing is trust. You can provide information but people will only take that information if they deem the source trustworthy. Some people do not trust the big pharmaceutical companies and will not take any information from them as credible. The population must be studied to know sources they are most likely to trust. PR can then use these agencies to promote vaccine acceptance. In Ghana, for example, most of the Christian population are more likely to accept information from their church leadership than the Government. [Official 002; 4 years’ experience]*.

However, certain pitfalls were discovered that must be taken into consideration when designing PR programmes.

> *However, failure will stem from frequent shortages of vaccines and lack of political will to implement programmes related to vaccines. [Official 010; 3 years’ experience]*

In addition, programmes must consider the high rates of illiteracy in rural areas and any potential conflicts with religious beliefs.

> *I will tell you about an incident that shocked all of us here. When school children returned to school after the COVID-19 hiatus, several school visits were planned…As soon as we parked the Service’s vehicle on the school compound, all the children trooped out and headed home. This was during the peak of debates about a team of French scientists purported to be planning vaccine trials in Africa. We had to go into the community to reassure the parents that no vaccination was planned. [Official 013; 10 years’ experience]*

A key lesson learnt here by the team was of the importance of community leaders in the planning of even routine public health exercises. After this incident, the district health team was able to gain the confidence of community leaders after they were thoroughly assured that no vaccination was planned.

> *“There was an occurrence where a person shared that they experienced an adverse event following vaccination (AEFI); within that period, the average number of people who came in for the vaccine decreased. Notwithstanding, it is observed that most people, having heard that someone they knew took the vaccine had the motivation to take the vaccine as well…we must get those who the people listen to so they get them to change*.*” [Official 004; 5 years’ experience]*.

The Ghana Health Service broadcast all messages in the local dialect; on radio for the cities and towns and on the local megaphones at the community level.

## Discussion

The present work successfully explored strategies used by health care professionals working with the GHS to address COVID-19 related vaccine hesitancy in the country through in-depth interviews with key informants. A thematic content analysis of the informant interviews confirms that healthcare workers perceive vaccine hesitancy to be a threat with the potential to undermine strategic goals of their organization especially related to the attainment of herd immunity against COVID-19. In response, a number of PR strategies including informative, facilitative, persuasive, coercive and motivational PR strategies were employed to diffuse this threat across both traditional and emerging media networks.

Deterioration of public trust in vaccination in general has been reported by many authors in line with the present study (10). General suspicion of vaccination, misunderstandings about infection severity, and an aversion for medical innovation are known to propel vaccine hesitant behaviour and even non-compliance to public health measures such as the wearing of nose masks (22). The consensus was that although the concept of herd immunity was an important one and had been communicated as an important organizational goal in the fight against the COVID-19 pandemic by credible and authoritative sources and therefore was well assimilated and aspired to, the communication had not been unambiguously passed on to the general public who in turn had conflicting explanations about how this goal could be achieved.

Healthcare workers are expected to play a significant role in addressing commonly held misconceptions and driving widespread acceptance for vaccine products among the public (23). The Centers for Disease Control and Prevention (CDC) highlights 12 strategies to promote vaccine confidence and uptake. These include, vaccine ambassadors, medical provider vaccine standardization, medical reminders, motivational interviewing, financial incentives, school-based vaccination programs, home-delivered vaccinations, workplace vaccination, effective messaging by trusted messengers, provider recommendation and combating misinformation (24). Consistent with this position, officials of the GHS demonstrated an awareness of their role in the fight against misinformation and maintaining positive vaccine behaviour among members of the public.

The role of health advocate requires employees to develop an awareness of key organizational goals and to be able to work towards achieving them. Here, this was done through various PR strategies and communication tools. Strategies commonly used involved information dissemination, facilitation of desired behaviour, persuasion, coercion, and motivation (25).

Organizations typically use the informative strategy as to advance objective facts about situations with the assumption of well-educated and self-motivated publics. Without necessarily making conclusions or advancing opinions, this strategy succeeds because the intended audience will derive the expected pattern of behaviour by making the right conclusions from available data provided it is accurate. A key function of this strategy is providing public education about subjects that inform or are hinged upon organizational goals. In the case under study, we see the GHS advancing knowledge about the vaccine manufacturing process, the relevance and conduct of trials and in general even about public health concepts such as herd immunity to counter anti-vaccine campaigns on social media.

Educative campaigns may offer new or alternative solutions to existing threats to an organization’s interests such as an under-informed audience. Additionally, they may stand out by the use of unbiassed language and organic communication about controversial subjects to aid comprehension. According to Werder (26) educative campaigns are most effective as a PR strategy when organizations use them consistently over a lengthy time period to drive behavioral change. This consistent approach makes the strategy more credible in the public sphere as foundations are laid for future learning. Additionally, they can be useful for stating a problem or threats to organizational goals and establishing confidence that once a problem is known, it can be resolved (27).

A facilitative strategy was accomplished by making resources available to a public that allow it to act in ways that it is already predisposed to act such as individuals who are already eager to be vaccinated. The study provides evidence that the GHS offered directions for members of the public to get easy access to mobile vaccination sites within communities. According to Zaltman (27), facilitative communication is useful when addressing challenges that belong to the shared consciousness of an organization and its public. In such cases, there is an agreement on the specific remedy to be implemented and there is openness to external assistance. Facilitative strategies may be an effective way for the GHS to promote greater public awareness for the existence of various support schemes for achieving both organizational and client goals. Also, the GHS may employ facilitative strategies to motivate individuals who are amenable to behavioral change because without the incentive, these persons may lack the resource capacity required to contribute to organizational goals.

In many ways the PR campaign followed by the GHS can be described as an exercise in persuasion. According to Werder (25), a persuasive strategy derives its significance by appealing to shared values or emotions in the external environment. This strategy as used by the GHS emphasized a tailored presentation of facts to invoke public sympathy and concerted action. Persuasive PR managers are noted for the use of language that is not objective but goes the extra mile to show the cross-cutting significance of a matter at hand to compel initiative. Persuasive strategies and communication used by the service are often directive, containing an implicit/explicit call to action. According to Zaltman (27), “persuasive strategies are useful in cases where threats to organizational goals are not evident or considered essential by an organization’s publics, and as a result, commitment falls below expectation because the proposed organizational solution is in doubt.” Usually persuasive public relations strategies complement other PR strategies very well in contexts where publics are expected to take voluntary steps and the organization has no control through the management of resources valued by the public such as salaries (26).

Officials also recognized that a high rate of illiteracy among the populace could be both a set-back and an advantage. Obviously, this high rate of illiteracy becomes a problem when developing persuasive PR strategy because for a health organization with rich expertise who could speak convincingly about technical issues, a public with a high need for cognition would be an advantage when developing a PR strategy. The fundamental idea of the elaboration likelihood model of persuasion is that when a group has a high appetite for mental exertion and crave cognition, the processing of communication will mostly take place along a central route. The desired changes in attitudes and behaviour follows on from deliberate and conscious processing of the arguments spelt out in the communication and as a result, such the newly informed behaviour is likely to be persistent (28). Future patterns of behaviour for individuals with an appetite for complex and detailed information are highly predictable as a result of the extensive elaboration (29, 30) and these individuals believe that they convinced themselves and were not convinced by the originator of the message. However, when faced with a large rural population and a high rate of illiteracy, it is safe to conclude that elaboration likelihood is low and the processing of communication must follow a peripheral route which requires little mental exertion. For these population groups, the GHS had to concentrate the communication using peripheral cues such as the credibility of the messenger and cues (29). It follows from these mechanisms that behaviours acquired through the latter route are mostly not hinged on the presentation of more superior quality arguments, are temporary, and are not as revealing of subsequent sentiment as those formed using the central route (29).

## Strengths and limitations

This report is the first to document experiences of using PR strategies to achieve organizational goals in the context of public health management in sub-Saharan Africa. The explorative approach facilitated a more nuanced appreciation of the phenomenon under investigation and exposed important themes. However, it also presented a number of limitations. For instance, it was impossible to quantify the exact contribution of individual strategies employed to the overall outcome. It was also unclear whether the findings were applicable to other vaccine scenarios beyond COVID-19 vaccines. More importantly, further studies would be required to clarify our understanding of potential confounding variables within the target population and the impact of global healthcare organizations such as the World Health Organization and the Global Alliance for Vaccines on the study’s findings.

## Conclusions

The multi-pronged PR strategies deployed by the GHS across both traditional and emerging/social media networks have been largely efficacious and beneficial in combating vaccine hesitancy. These were a combination of informative, facilitative, persuasive, motivational and coercive PR strategies. The GHS will likely benefit from entrenching these PR strategies into an institutional culture and sustaining same in order to guarantee positive behaviour towards vaccination in Ghana in the future. The strategies are recommended for healthcare managers to prevent vaccine hesitancy in similar contexts. While these PR strategies may be useful for improving vaccine behaviour, or at least forestalling a crisis of vaccine hesitancy, they may also be applicable to other organizational goals. Quantitative research approaches are required to determine the extent to which these PR strategies are practiced and correlated to the attainment of organizational targets.

## Data Availability

All data generated or analyzed during this study are included in this published article and its supplementary information files (S2 Appendix B.xlsx: Data file).

## Abbreviations

AIDS: Acquired Immune Deficiency Syndrome
CIM: Chartered Institute of Marketing
CIPR: Chartered Institute of Public Relations
COREQ: Consolidated criteria for reporting qualitative research
COVID-19: Corona virus disease
EI: Executive instrument
ELM: Elaborated Likelihood Model
GHS: Ghana Health Service
HIV: Human immunodeficiency virus
ICT: Information and Communication Technology
MOH: Ministry of Health
MP: Member of Parliament
NFC: Need for cognition
PNDCL: Provisional National Defence Council Law
PR: Public relations
SARS CoV: Severe acute respiratory syndrome coronavirus
WHO: World Health Organization

## Declarations

### Ethics approval and consent to participate

Ethical approval was obtained from the Committee for Human Ethics, UENR. All participants documented voluntary consent for data provided to be disseminated. All personal records of participants were anonymized.

### Consent for publication

Not applicable

### Authors’ contributions

**RM, DDA and ETD (author 3) are** credited with conceptualizing the study, data transcription, analysis and writing the foundational draft of the manuscript. **EO and IWB** Contributed conceptually to the paper, reviewed, effected changes and provided feedback on all drafts and provided approval for submission. **KOB** Contributed conceptually to the manuscript, participated in data analysis, provided feedback on all drafts and gave approval for submission. **ETD (author 5)** Contributed conceptually to the paper, effected changes to and provided feedback on all drafts and gave approval for submission.

## Acknowledgements

The authors are grateful to the Ghana Health Service for providing reliable feedback through their officers and to all officials who graciously granted interviews.

## Supplementary files

S1 Appendix A.docx: Interview guide for in-depth interviews

S2 Appendix B.xlsx: Data file

